# Anthocyanin-rich extract mitigates the contribution of the pathobiont genus *Haemophilus* in mild-to-moderate ulcerative colitis patients

**DOI:** 10.1101/2024.09.30.24314597

**Authors:** Yannik Zobrist, Michael Doulberis, Luc Biedermann, Gabriel E Leventhal, Gerhard Rogler

**Author notes:** Address correspondence to: Prof. Rogler Gerhard, Chief of Department of Gastroenterology and Hepatology, Department of Medicine, Zurich University Hospital, Zurich 8091, Switzerland.

## Abstract

**Objective:** Anthocyanins (AC) have been shown to elicit anti-inflammatory and antioxidant effects in several animal models of colitis. Furthermore, AC lowered biochemical disease activity in our double-blind randomized trial in patients with ulcerative colitis. Here, we report on the changes in the faecal microbiome composition in the patient upon AC exposure in this trial.

**Methods:** Ulcerative colitis patients received a 3g daily dose of an AC-rich bilberry extract (ACRE) for eight weeks. We determined the microbiome composition in longitudinal stool samples from 24 patients and quantified the degree of change over time. We also correlated the relative abundances of individual microbial taxa at different time points to faecal concentration measurements of calprotectin.

**Results:** Microbiome compositions did not change over time as a result of the intervention, both in terms of alpha and beta diversity. Before the intervention, Haemophilus parainfluenzae was positively correlated with faecal calprotectin concentrations, and this correlation persisted in placebo-treated subjects throughout the study. In contrast, the correlation between H. parainfluenzae and the concentration of faecal calprotectin vanished in ACRE-treated subjects, while the relative abundance of H. parainfluenzaae did not change. Conclusion: Our results suggest that ACRE treatment mitigates the contribution of H. parainfluenzae to inflammation. Further research is warranted to better comprehend the role of microbial composition in response to medical therapy including AC-rich extract in ulcerative colitis patients.

**Key messages:** *1. What is already known on this topic:* Anthocyanins are a promising therapeutic alternative for ulcerative colitis patients given their effect on faecal calprotectin, excellent safety profile, and low costs.

*2. What this study adds:* The administration of anthocyanin-rich extract to ulcerative colitis patients in a Phase 2a study reduced faecal calprotectin concentrations by mitigating the contribution from the pathobiont genus Haemophilus but did not affect the overall microbiome composition.

*3. How this study might affect research, practice or policy:* Our results could aid in targeted treatment of ulcerative colitis patients with anthocyanins, according to individual microbial composition.

## Introduction

The incidence and prevalence of inflammatory bowel disease (IBD) and in particular ulcerative colitis (UC) is rising worldwide.[1] The exact pathophysiology of UC remains unknown, with various factors thought to influence the emergence of UC,[2] complicating early detection in cases of suspected IBD, the current non-invasive diagnostic gold standard is determination of faecal calprotectin. The latter, represents a crucial non-degradable leukocyte protein with the best correlation to endoscopic inflammatory indices compared to CRP. It effectively distinguishes among mild, moderate and severe inflammation.[3]

Approximately two-thirds of UC patients with mild-to-moderate disease activity can be successfully treated with the anti-inflammatory drug mesalamine (5-ASA). However, non- responders to 5-ASA treatment remain a clinical challenge, with severe cases requiring invasive treatments such as colectomies. Although several other potentially effective medications exist for UC, their typical safety profile is far from ideal, with substantial short and long-term toxicity risks and with high annual costs for newer treatment options such as biologics.[4] Because of this, there is strong interest from IBD patients for complementary therapeutic options that are perceived as “natural and holistic” with fewer side effects.[5]

Anthocyanins (AC) a specific type of deglycosylated anthocyanidins, are an alternative herbal remedy for UC. ACs are concentrated in various vegetables and berries, particularly bilberries,[6,7] and are known to have antioxidant and anti-inflammatory effects[7,8] and have thus been preclinically investigated for the treatment of UC. Animal model studies have shown that AC treatment has beneficial effects on dextran sodium sulphate (DSS)-induced colitis. These effects include lower histological scores, reduced cytokine release, less colonic shortening (fibrosis), less weight loss, less hepatosplenomegaly, increased intestinal permeability (which favours bacterial translocation), and lower abundances of pathogenic bacteria compared to control groups.[9,10]

Mechanistically, it is feasible that ACs can act on the microbiome in the colon. The majority of ingested ACs bypass absorption in the stomach and small intestine and reach the microbiome-rich colon. There, certain bacteria deglycosylase and metabolize ACs into phenolic acids such as protocatechuic, vanillic, syringic, gallic, or 4-hydroxybenzoic acid.[11] These bacterial fermentation derivates exhibit beneficial chemoprotective effects due to their intrinsic anti-inflammatory and antioxidative properties.[12] Interestingly, positive effects of AC such as increased short chain fatty acids (SCFA), reduced spleen weight, re-extension of colon length, or ameliorated histological scores could not be achieved or replicated in germ-free mice,[9] suggesting a key role of the microbiome in the AC mode of action Intake of AC also alters the intestinal microbial composition,[10,13] However, it remains unclear whether the observed effects of AC on the human gut microbiome are direct, or indirect via a modulation of the microbial composition and the microbiome’s metabolic properties.

We recently reported on a multicentre, double-blind, randomized, placebo-controlled phase IIa study to confirm a therapeutic effect of ‘AC rich extract’ (ACRE) therapy previously observed in a pilot study.[14] In the phase IIa study, we observed that faecal calprotectin concentrations decreased during the treatment phase and subsequently increased again after stopping ACRE therapy.[15] Here, we asked whether the effect of ACRE on faecal calprotectin was mediated by the microbiome. We hypothesized that the effect acted on the microbiome either directly, by inhibiting or promoting certain bacteria, or indirectly, by modulating potential negative effects of certain microbiota. To answer this, we characterized the bacterial microbiome composition in the UC patients throughout the study and particularly investigated whether specific genera might be associated with the observed effect of faecal calprotectin reduction during the intervention.

## Methods

### Ethics

The study was carried out in accordance with principles enunciated in the current version of the Declaration of Helsinki,[16] the guidelines of Good Clinical Practice[17] issued by International Council for Harmonization of Technical Requirements for Pharmaceuticals for Human Use and Swiss regulatory authority’s requirements. The project was approved by the Ethics Committee Zurich (BASEC-Nr, 2017-00156).

### Study design

The full protocol and study design can be found in the supplementary files. Briefly, we included individuals over 18 years of age who had a UC diagnosis for at least three months with a modified Mayo score of 6-12 and a disease activity despite therapy with 5-ASA and steroids. A detailed description of all inclusion and exclusion criteria can be found in the appendix (Supplementary Table 1).

Study participants were randomly assigned to either the verum or placebo group in a 2:1 ratio. The verum group received a daily dosage of three times 2x500mg of an ACRE provided by Walther Riemer GmbH (Nimbo Green, China), corresponding to 100g dried bilberries or 840mg of anthocyanins per day. The screening phase spanned four weeks, followed by an eight-week intervention period and a subsequent three-week follow-up phase. Three consultations were conducted during the intervention, with stool samples and other relevant data collected at all time points. A detailed study procedure description is available in the Appendix (Supplementary 2).

### Pre-analytics

The stool was collected with an OMNIgene® GUT kit (DNA Genotek Inc.) and later stored at -20°C until DNA extraction and sequencing (performed by Microsynth AG, Balgach, Switzerland). Amplicon sequencing was performed using Illumina MiSeq paired-ends sequencing technology. The hypervariable region V4 of bacterial 16S rRNA genes was amplified using the primers 515F (GTGCCAGCMGCCGCGGTAA), and 806R (GGACTACHVGGGTWTCTAAT) to generate an approximate amplicon size of 300bp. Multiplexed libraries were constructed with help of the two-step PCR protocol (as recommended by Illumina). The paired-end reads were trimmed of their locus specific primers and subsequently merged to in-silico reconstruct the amplified region resulting in a total of 22.74 million reads, from which Amplicon Sequence Variants (ASVs) were then identified.

ASVs were inferred using Dada2 v1.18.0 with read length filtering set to 250 and 210 and maximum expected errors (maxEE) set to 4 and 5 for forward and reverse reads, respectively, and inference performed in “pseudo pool” mode. Read pairs were merged with minimum overlap (minOverlap) of 20, and bimeras were removed using the consensus method. The prepared GTDB r95 taxonomic database for Dada2 (GTDB ref, Dataset Ref) was used for taxonomic annotations via the assignTaxonomy function in Dada2.

### Statistics

We obtained ASVs from 35 subjects at different study time points. After excluding data from individuals lacking baseline calprotectin measurements or with data available at fewer than 3 study time points, our final cohort comprised of 24 patients, with 17 in the verum group and 7 in the placebo group.

Genus-level relative abundances were computed by summing the counts of all ASVs that were assigned to a specific genus and normalizing by all reads. Whenever log-transformed relative abundances were computed, a pseudo count of 1 was added to the read counts.

The Shannon diversity, H, was computed at the relevant level of taxonomic resolution (e.g., genus). Subsequently, resulting effective number of genera was computed as the exponential of the Shannon diversity, exp(H), and represents the equivalent number of evenly distributed taxa with the same Shannon diversity. Comparisons between treatment groups were performed for each timepoint individually using Wilcoxon signed-rank tests.

We computed the Aitchison distance, i.e. the magnitude of change in microbiome composition between the centre-log-ratio transformed relative abundances. Comparisons between groups or timepoints were performed using Wilcoxon signed-rank tests.

The change of individual taxa (e.g., genera) over time following baseline was estimated from a linear mixed model of the log10-transformed relative abundances with treatment as a fixed effect and by treating timepoint as an integer including a random slope for the effect of time and a random intercept for each individual.

To quantify the association of individual microbiome genera with calprotectin levels prior to the intervention, we performed individual linear mixed model regressions of the log_10_-transformed calprotectin concentrations on the log_10_-transformed relative abundances for those genera that were detected in at least 50% of the samples. To capture potential variability in the microbiome over time, we used both the screening and baseline abundances and accounted for subject identity as a random effect. We adjusted the p-values using the method of Benjamini-Hochberg.

## Results

We previously reported that ACRE treatment reduced faecal calprotectin concentrations resulting in a significantly lower concentration in the ACRE group compared to the placebo group by the third visit,[15] but not at earlier visits. In order to better understand the dynamics of how the faecal calprotectin over time, we estimated a per- subject change in faecal calprotectin using a linear mixed model. Faecal calprotectin decreased steadily during treatment in the ACRE group (p = 0.0243), but did not change in the placebo-treated patients (p = 0.427; Supplementary Figure 1). The concentration subsequently increased again following the end of the treatment. This suggests that ACRE treatment might act gradually and consistently.

A gradual action of ACRE on the microbiome might manifest in different ways. First, in a direct way, ACRE treatment might gradually shift the microbiome composition over time away from a pro-inflammatory state. Second, in an indirect way, ACRE treatment might not affect the microbiome composition per se, but instead mitigate any pro-inflammatory impact of the microbiome. In the first case, we would expect the microbiome composition to gradually change during ACRE treatment—but not during placebo treatment, and in the second case we would expect the microbiome to not change in either group. We did not observe any evidence of a systematic change in microbiome composition in either treatment group. We quantified the change in microbiome composition in terms of alpha and beta diversity. For alpha diversity, we modelled the change in the effective number of genera using the same linear mixed model approach as for faecal calprotectin with a treatment- specific slope and common intercept at baseline, and per-subject random slopes and intercepts. Alpha diversity did not change over time in either treatment group (Figure 1a). For beta diversity, we quantified the degree of change in terms of the Aitchison distance between Baseline and Visit3 for each subject, and compared this to the distribution of distances between subjects at Baseline. The Aitchison distance between Baseline and Visit3 was not significantly different in the ACRE and placebo groups (p = 0.697, Figure 1c), implying that individual patients mostly retained their microbiome composition during treatment (Figure 1b). Taken together, we do not find evidence that ACRE treatment modifies the microbiome composition directly.

**Figure 1.**
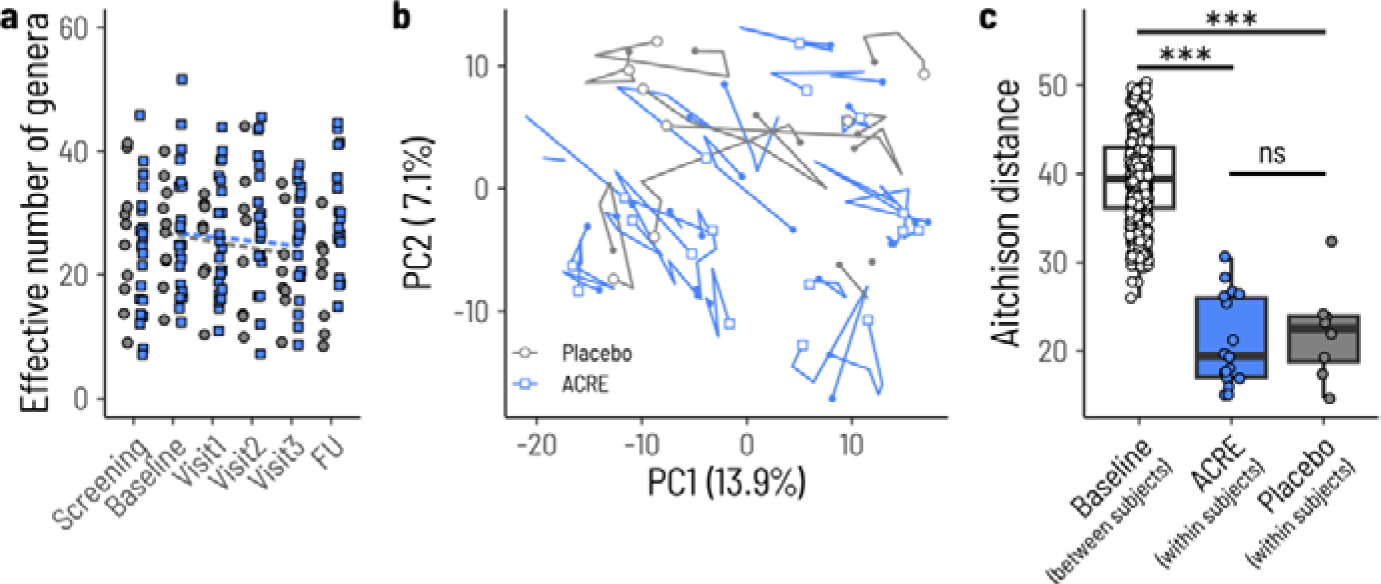
Microbiota composition is not affected by ACRE treatment. a. Microbiome alpha diversity does not change during treatment. Blue squares and grey circles show the alpha diversities at different timepoints in the study for the ACRE and placebo groups, respectively. Symbols from the same patients are connected by lines. The dashed lines show the estimated slopes from a linear mixed effects model that are not significantly different from zero (p = 0.260 and p = 0.215). b. Principal component analysis (PCA) of the faecal microbiota based on genus consumption over time. Each path represents an individual patient. (baseline: filled shapes, follow-up: open shapes). c. Aitchison distances between subjects at baseline, or between Baseline and Visit3 for the Bilberry and Placebo groups. ns = non- significant, *** = p value < 0.00005.

Given that ACRE treatment does not modify microbial composition directly, we next turn to the second option where ACRE acts indirectly, implying that there are certain microbiota characteristics that promote inflammation that are then mitigated by ACRE treatment. To identify such characteristics, we analysed whether certain genera correlated with faecal calprotectin concentrations before the start of the intervention. To increase statistical power, we grouped the screening and baseline samples together but accounted for subject identity as a random effect.

We identified 13 genera whose relative abundances were either positively or negatively associated with calprotectin levels prior to the intervention (Figure 2a). Of these, only two—Haemophilus and Parasutterella—remained significant after correction for multiple testing at a false-discovery rate below 0.1 (Supplementary Table 3). Increasing relative abundances of the genus Haemophilus were associated with higher concentrations of faecal calprotectin (p_adj_ = 0.053, conditional R^2^ = 0.43), whereas higher relative abundances of Parasutterella were associated with lower concentrations of calprotectin (p_adj_ = 0.053, conditional R^2^ = 0.52) (Figure 2b).

**Figure 2.**
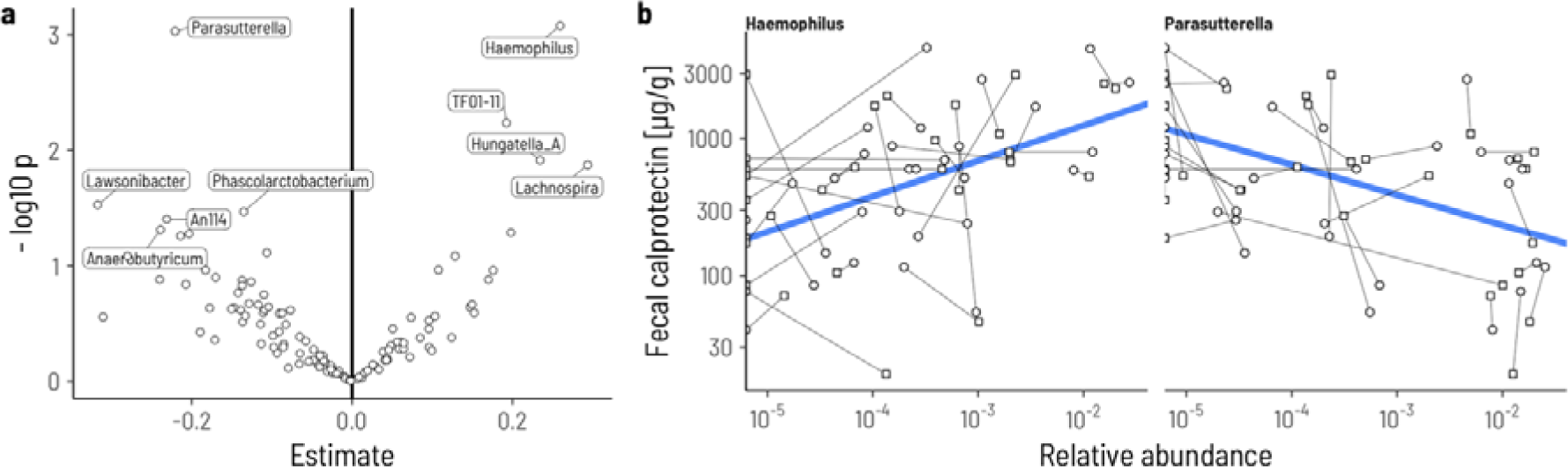
Genera associated with high/low fecal calprotectin concentration prior to the intervention. a. Volcano plot of the regression slopes and their individual p-values. Genera with p < 0.05 are labelled. b. Visualization of the relationship between relative abundance (x-axis) and fecal calprotectin concentrations for the two genera with a false discovery rate < 0.1. Samples from the same individual at Screening (circles) and Baseline (squares) are connected with a line.

This analysis identifies Haemophilus abundance as a potential contributor to inflammation and conversely Parasutterella as a potential mitigator of inflammation in the subjects before initiating treatment. We presumed that if these contributions were robust, then they should persist throughout the study in the placebo-treated individuals in which we do not expect any modulation of the interaction between microbiome and inflammation.

The association of Haemophilus and faecal calprotectin persisted in the placebo group throughout the intervention, while the association for Parasutterella did not. We used all sampling points after the start of the intervention, i.e., visits 1-3 and follow-up and accounted for subject identity as a random intercept. Haemophilus relative abundance remained significantly correlated with faecal calprotectin concentrations in the placebo group after the start of the intervention (p = 0.0347, conditional R = 0.367, Supplementary Figure 2). In contrast, the negative correlation between Parasutterella Relative abundance and faecal calprotectin was not significant after the start of the intervention (p = 0.673, conditional R = 0.28). This suggests that Haemophilus might indeed contribute directly to inflammation.

We hypothesized that if ACRE directly exerts an influence on the microbiota, for instance by depleting Haemophilus abundance, then a correlation between Haemophilus and faecal calprotectin would be expected to remain in the ACRE group, with a potential overall decrease in Haemophilus abundance. Alternatively, if ACRE were to modify the mechanism with which Haemophilus contributes to inflammation, then we would not expect an effect on Haemophilus abundance but rather that there would nevertheless be a decrease in faecal calprotectin concentration.

To test these hypotheses, we first investigated whether the relative abundance of Haemophilus changed during ACRE treatment. Second, we investigated whether the relationship between the abundance of Haemophilus and faecal calprotectin concentration remained in the ACRE group after the start of the intervention.

ACRE treatment did not impact Haemophilus abundance but did negate the association between Haemophilus and faecal calprotectin. The estimated change in Haemophilus abundance over time in the ACRE group was not significantly different from zero (p = 0.345, Figure 3). However, faecal calprotectin was no longer significantly associated with Haemophilus relative abundance after the start of the intervention (p = 0.422, conditional R^2^ = 0.42) (Figure 4). Closer inspection of the ASVs that were identified as Haemophilus in our data revealed only a single ASV that mapped with 100% identity to the strain Haemophilus parainfluenzae ATCC 33392. Taken together, these results indicate that ACRE treatment modifies the interaction between Haemophilus parainfluenzae and inflammation, rather than by directly decreasing Haemophilus abundance.

**Figure 3.**
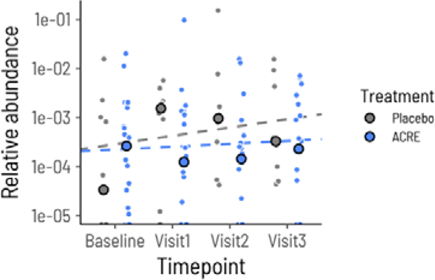
Haemophilus relative abundance remains stable during treatment. Each small circle shows the relative abundance of the genus Haemophilus in either the placebo group (grey) or ACRE group (blue). The large circles show the mean within the groups for each timepoint. The dashed lines show the estimated regression slope of log10 relative abundance over time.

**Figure 4.**
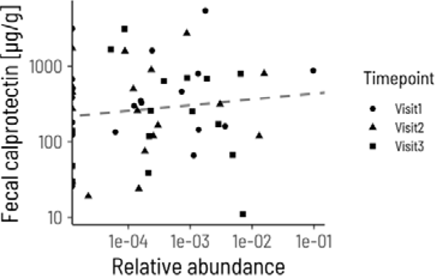
Haemophilus relative abundance is not significantly correlated with faecal calprotectin during ACRE treatment. Each point corresponds to a sample from a patient at visit 1 (circles), visit 2 (triangles), or visit 3 (squares). The dashed line shows the estimated regression slope from a linear mixed model with subject as a random effect.

## Discussion

In our study we aimed to investigate to what degree the previously observed effect of ACRE treatment on lowering faecal calprotectin concentrations in a clinical study in subjects with mild to moderate UC was linked to a modulation of the microbiome in these subjects. . Unexpectedly, we did not observe any substantial shifts in microbiome composition induced by ACRE treatment. Instead, we observed that ACRE treatment mitigated the association between the abundance of the genus Haemophilus in the faecal microbiome and the faecal calprotectin concentration. Prior to initiating the study, patients with higher abundances of Haemophilus also had higher concentrations of faecal calprotectin and this correlation persisted in the placebo-treated patients. In contrast, the correlation vanished during ACRE treatment. Therefore, ACRE administration did not reduce faecal calprotectin concentrations by directly modulating the microbial composition, but rather by indirectly affecting the proinflammatory mechanisms of bacterial taxa such as Haemophilus.

The observation that treatment with AC did not affect the microbiome composition is unexpected based on previous mouse studies in the literature of AC impacting microbiome alpha and beta diversity in UC. Two studies observed an increased Shannon index and a significant beta diversity shift after ingesting 200mg/kg AC for 7 or 17 days, respectively.[10,18] Another four-week study with daily intake of 3.47mg suggested a potential increase in alpha diversity post-anthocyanin intake.[19] However, these studies were performed in mice with DSS-induced colitis, used sources of AC other than bilberries that may impact the effect,[20] and had higher daily doses per kg body weight (173- 200mg/kg) compared to our study (ca. 11.8mg/kg; assuming an average European weight of 71kg).

The genus Haemophilus has previously been reported to be increased in abundance in IBD patients.[21–24] The positive correlation we observed between the relative abundance of Haemophilus and faecal calprotectin concentrations is in line with a previous work that found an association of Haemophilus with more severe disease.[25] Consistent with our results, two studies have reported an increased presence of Haemophilus spp. in UC patients that is particularly pronounced in active stages.[26,27] Thus, emerging evidence suggests that Haemophilus spp. may be considered as a potentially pathogenic genus for UC. Moreover, studies have demonstrated that Haemophilus parainfluenza exhibits strong IgA coating in individuals with UC.[28] IgA, the predominant dimeric antibody in the intestinal mucosa, plays a crucial role in coating and neutralizing pathogens. IgA coating is also observed in endogenous microbiota, though to a lesser extent than with pathogens.[29] Given that certain genera influencing UC severity share similarities with pathogens,[30] Palm et al. and Shapiro et al. suggest pathogenic microbiota may be more heavily coated with IgA.[31,32] Furthermore, more active UC is associated with increased IgA coating and higher faecal IgA levels[33] and on the other hand mice with elevated IgA levels and therefore higher amount of coated bacteria demonstrate greater resistance to DSS-induced murine colitis.[34] Finally, blueberry ingestion is linked to increased IgA secretion.[35,36] In summary, it is reasonable to hypothesize that IgA secretion is increased by higher inflammatory activity or pathogenic microbes, thus coating the more pathogenically active microbiota trying to provide control of colitis. AC, in turn, seem to facilitate IgA production, exerting their potentially anti-inflammatory effect on UC. ACRE may stimulate IgA secretion resulting in a more intensive coating of pathobiont Haemophilus, thereby reducing its proinflammatory potential. However, other potential anti-inflammatory mechanisms of ACRE via the microbiome are conceivable, such as promoting beneficial SCFA production, reducing colonic shortening, or enhancing epithelial barriers.[9,13]

The observation of a clinically meaningful decrease in a key biochemical parameter that is linked to disease activity in UC suggests that AC might be considered as a viable treatment option in UC that warrants further evaluation. While it is unlikely that anthocyanins exert their effects exclusively by mitigating the proinflammatory potential of Haemophilus spp., their potential benefits may be more pronounced in individuals with higher Haemophilus spp. abundance in their intestinal microbiome. Additionally, probiotic therapies might prove valuable in reducing pathobionts such as Haemophilus spp. in UC patients, fostering a more favourable microbial microenvironment for the disease. Nevertheless, larger studies are essential to reach accumulation of positive evidence and secondarily a consensus on the composition of the microbiota and its analysis in order to take a step towards personalized medicine, especially with complementary options such as ACRE.

A particular strength of our study is the randomized double-blind design with a novel intervention among the first to investigate the impact of a high-dose anthocyanin intervention on the microbiome composition in UC patients. While there are already some investigations on this topic in animal models,[9,10,13,18–20] this is to the best of our knowledge the first human data. Another strength of our study is that we repeated assessments of the key parameters, microbiome composition and faecal calprotectin concentration, over time for the same subjects that enables us to account for some of the intra-individual variability.

Several limitations of our study have to be acknowledged. Firstly, our sample size comprising of only 24 patients is rather small and for this reason we have taken care that our statistical inferences are appropriate A further limitation that is shared by most other microbiome studies is that our work considers only relative abundances of microbial taxa and focuses primarily on faecal calprotectin. We did not investigate other related parameters such as the microbial metabolome or immunologic factors like intestinal IgA production. Finally, we exclusively focused on bacterial taxa of the intestinal microbiota and did not take other microbes into account, such as fungi, viruses, or protozoa.

In conclusion, we provide evidence that the anti-inflammatory effect of the ACRE intervention was not mediated via a relevant modulation of the microbiome, as originally thought, but rather via a regulation of the pro-inflammatory effect of its pathobionts such as Haemophilus parainfluenzae. Whether this effect is directly caused by the administered AC or rather by their degradation products after metabolization by the microbiome remains unclear. Regarding the intake of AC, the exact interaction of anthocyanins and the microbiome in UC has not yet been clarified and remains vague. Which biochemical and physiological links are involved and what might be the overall effect on the disease needs to be further investigated.

## Data Availability

All data produced in the present study are available upon reasonable request to the authors

## Acknowledgements

The authors wish to thank all the patients for their cooperation as well as their families for their support during this endeavour.

## Ethical Considerations

### Financial disclosures and conflicts of interest

MD reports traveling fees from Takeda, FALK, Abbvie as well as consulting fees from Takeda LB reports fees for consulting/advisory board from Abbvie, MSD, Vifor, Falk, Esocap, Calypso, Ferring, Pfizer, Shire, Takeda, Janssen, Ewopharma. GR declares consulting fees from Abbvie, Augurix, BMS, Boehringer, Calypso, Celgene, FALK, Ferring, Fisher, Genentech, Gilead, Janssen, MSD, Novartis, Pfizer, Phadia, Roche, UCB, Takeda, Tillots, Vifor, Vital Solutions and Zeller; speaker’s honoraria from Astra Zeneca, Abbvie, FALK, Janssen, MSD, Pfizer, Phadia, Takeda, Tillots, UCB, Vifor and Zeller; and grants support from Abbvie, Ardeypharm, Augurix, Calypso, FALK, Flamentera, MSD, Novartis, Pfizer, Roche, Takeda, Tillots, UCB, and Zeller. GEL is an employee and shareholder of PharmaBiome. All other authors declare no conflicts of interest

## List of Abbreviations

AC: Anthocyanin
ACRE: Anthocyanin Rich Extract
ASV: Amplicon Sequence Variants
CRP: C Reactive Protein
DSS: Dextran Sulphate Sodium
GTDB: Genome Taxonomy Database
IBD: Inflammatory Bowel Disease
IgA: Immunoglobulin A
PCA: Principal Component Analysis
PCR: Polymerase Chain Reaction
SCFA: Short Chain Fatty Acids
UC: Ulcerative Colitis
5-ASA: 5-Aminosalicylic Acid

## Supplementary Tables

**Supplementary Table 1:**
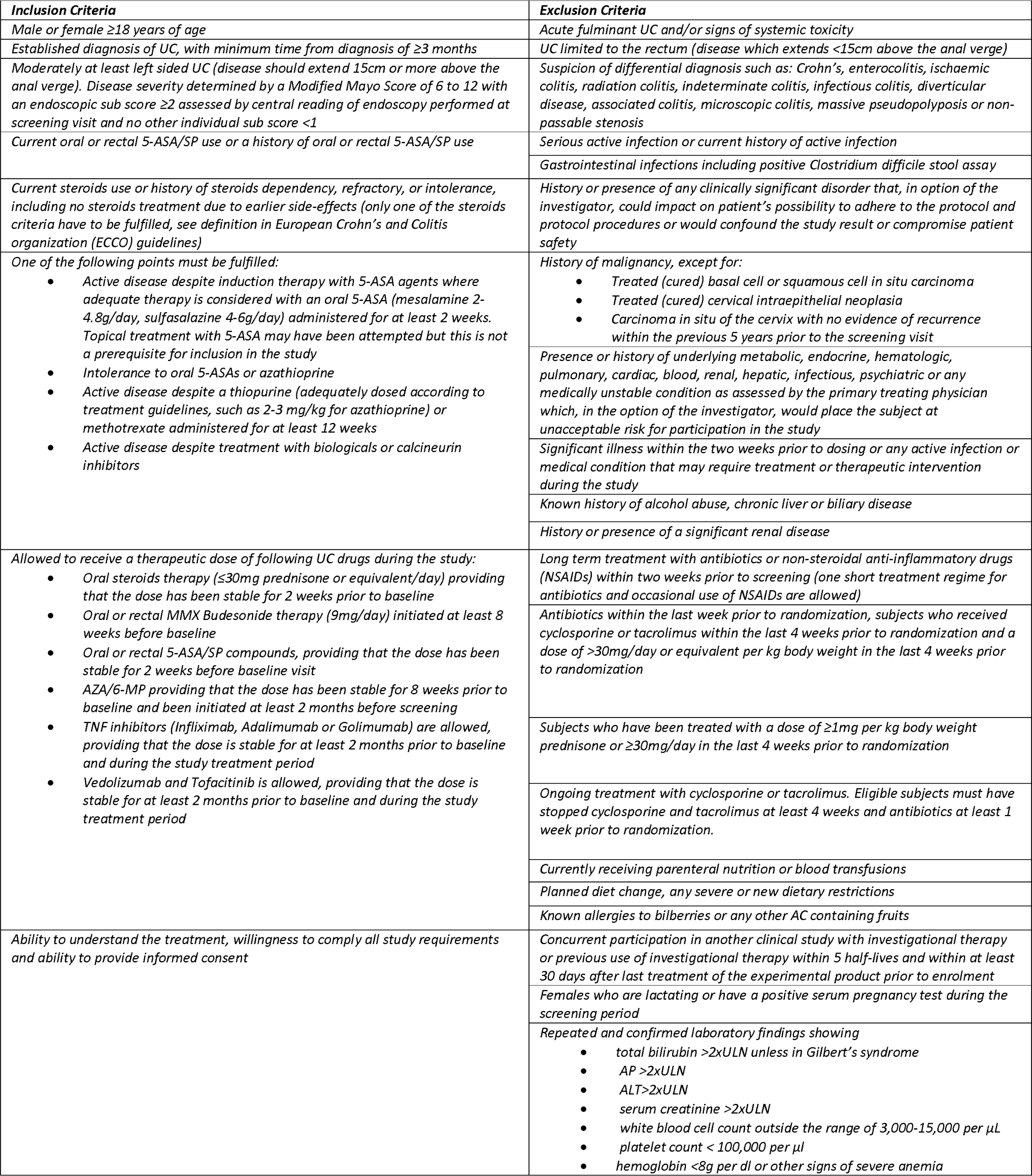
Detailled inclusion and exclusion criteria.

| **Inclusion Criteria** | **Exclusion Criteria** |
| --- | --- |
| Male or female $\geq 18$ years of age | Acute fulminant UC and/or signs of systemic toxicity |
| Established diagnosis of UC, with minimum time from diagnosis of $\geq 3$ months | UC limited to the rectum (disease which extends $< 15$ cm above the anal verge) |
| Moderately at least left sided UC (disease should extend 15 cm or more above the anal verge). Disease severity determined by a Modified Mayo Score of 6 to 12 with an endoscopic sub score $\geq 2$ assessed by central reading of endoscopy performed at screening visit and no other individual sub score $< 1$ | Suspicion of differential diagnosis such as: Crohn's, enterocolitis, ischaemic colitis, radiation colitis, indeterminate colitis, infectious colitis, diverticular disease, associated colitis, microscopic colitis, massive pseudopolypsis or non-passable stenosis |
| Current oral or rectal 5-ASA/SP use or a history of oral or rectal 5-ASA/SP use | Serious active infection or current history of active infection |
|  | Gastrointestinal infections including positive *Clostridium difficile* stool assay |
| Current steroids use or history of steroids dependency, refractory, or intolerance, including no steroids treatment due to earlier side-effects (only one of the steroids criteria have to be fulfilled, see definition in European Crohn's and Colitis organization (ECCO) guidelines) | History or presence of any clinically significant disorder that, in option of the investigator, could impact on patient's possibility to adhere to the protocol and protocol procedures or would confound the study result or compromise patient safety |
| One of the following points must be fulfilled: | History of malignancy, except for: |
| Active disease despite induction therapy with 5-ASA agents where adequate therapy is considered with an oral 5-ASA (mesalamine 2-4.8g/day, sulfasalazine 4-6g/day) administered for at least 2 weeks. Topical treatment with 5-ASA may have been attempted but this is not a prerequisite for inclusion in the study<br>Intolerance to oral 5-ASAs or azathioprine<br>Active disease despite a thiopurine (adequately dosed according to treatment guidelines, such as 2-3 mg/kg for azathioprine) or methotrexate administered for at least 12 weeks<br>Active disease despite treatment with biologicals or calcineurin inhibitors | Treated (cured) basal cell or squamous cell in situ carcinoma<br>Treated (cured) cervical intraepithelial neoplasia<br>Carcinoma in situ of the cervix with no evidence of recurrence within the previous 5 years prior to the screening visit |
|  | Presence or history of underlying metabolic, endocrine, hematologic, pulmonary, cardiac, blood, renal, hepatic, infectious, psychiatric or any medically unstable condition as assessed by the primary treating physician which, in the option of the investigator, would place the subject at unacceptable risk for participation in the study |
|  | Significant illness within the two weeks prior to dosing or any active infection or medical condition that may require treatment or therapeutic intervention during the study |
|  | Known history of alcohol abuse, chronic liver or biliary disease |
|  | History or presence of a significant renal disease |
| Allowed to receive a therapeutic dose of following UC drugs during the study: | Long term treatment with antibiotics or non-steroidal anti-inflammatory drugs (NSAIDs) within two weeks prior to screening (one short treatment regime for antibiotics and occasional use of NSAIDs are allowed) |
| Oral steroids therapy ($\leq 30$ mg prednisone or equivalent/day) providing that the dose has been stable for 2 weeks prior to baseline<br>Oral or rectal MMX Budesonide therapy (9 mg/day) initiated at least 8 weeks before baseline<br>Oral or rectal 5-ASA/SP compounds, providing that the dose has been stable for 2 weeks before baseline visit<br>AZA/6-MP providing that the dose has been stable for 8 weeks prior to baseline and been initiated at least 2 months before screening<br>TNF inhibitors (Infliximab, Adalimumab or Golimumab) are allowed, providing that the dose is stable for at least 2 months prior to baseline and during the study treatment period<br>Vedolizumab and Tofacitinib is allowed, providing that the dose is stable for at least 2 months prior to baseline and during the study treatment period | Antibiotics within the last week prior to randomization, subjects who received cyclosporine or tacrolimus within the last 4 weeks prior to randomization and a dose of $> 30$ mg/day or equivalent per kg body weight in the last 4 weeks prior to randomization |
| | Subjects who have been treated with a dose of $\geq 1$ mg per kg body weight prednisone or $\geq 30$ mg/day in the last 4 weeks prior to randomization |
|  | Ongoing treatment with cyclosporine or tacrolimus. Eligible subjects must have stopped cyclosporine and tacrolimus at least 4 weeks and antibiotics at least 1 week prior to randomization. |
|  | Currently receiving parenteral nutrition or blood transfusions |
|  | Planned diet change, any severe or new dietary restrictions |
|  | Known allergies to bilberries or any other AC containing fruits |
| Ability to understand the treatment, willingness to comply all study requirements and ability to provide informed consent | Concurrent participation in another clinical study with investigational therapy or previous use of investigational therapy within 5 half-lives and within at least 30 days after last treatment of the experimental product prior to enrolment |
|  | Females who are lactating or have a positive serum pregnancy test during the screening period |
|  | Repeated and confirmed laboratory findings showing<br>total bilirubin $> 2 \times \text{ULN}$ unless in Gilbert's syndrome<br>AP $> 2 \times \text{ULN}$<br>ALT $> 2 \times \text{ULN}$<br>serum creatinine $> 2 \times \text{ULN}$<br>white blood cell count outside the range of 3,000-15,000 per $\mu\text{L}$<br>platelet count $< 100,000$ per $\mu\text{L}$<br>hemoglobin $< 8$ g per dL or other signs of severe anemia |

**Supplementary Table 2:** Detailed study procedure description.

| **Study Periods** | **Screening** | **Treatment Period** |  |  |  | **Follow-up** |
| --- | --- | --- | --- | --- | --- | --- |
| **Visit** | **S** | **B** | **1** | **2** | **3** | **FUP** |
| **Day** | **-28 to -0** | **1 =**<br>**Baseline** | **15± 4d** | **28± 4d** | **56± 4d** | **86± 4d** |
| Subject Information and Informed Consent | x |  |  |  |  |  |
| Demographics | x |  |  |  |  |  |
| Medical History | x |  |  |  |  |  |
| Update Medical History |  | x | x | x | x | x |
| In- /Exclusion Criteria | x | x |  |  |  |  |
| Physical Examination | x | x | x | x | x | x |
| Vital Signs (pulse, blood pressure, temperature), Body Weight, Body Height<sup>1</sup> | x | x | x | x | x | x |
| Serum Laboratory Tests (Safety Lab) 2 | x | x | x | x | x | x |
| Pregnancy Test<sup>3</sup> | x | x | x | x | x | x |
| Urine analysis | x | x | x | x | x | x |
| Fecal Samples, stool bacteriology, C. diff. | x |  |  |  |  |  |
| Fecal Calprotectin | x | x | x | x | x | x |
| Microbiota Analyses | x | x | x | x | x | x |
| Resting-ECG | x |  |  |  |  |  |
| Sigmoidoscopy with 6 Biopsies and Endoscopic Mayo Subcore and Histology (Geboes Index) Recording for central reading | x |  |  |  | x |  |
| Complete Mayo Score | x |  |  |  | x |  |
| Partial Mayo Score (including PRO2) | x | x | x | x | x | x |
| Dispense Subject Diaries<sup>4</sup> | x | x | x | x | x |  |
| Collect Subject Diaries<sup>4</sup> |  | x | x | x | x | x |
| Assess quality of life (SIBDQ<sup>4</sup>, EuroQoL5D) | x | x | x | x | x | x |
| Distribution and/ or collecting of Study Medication |  | x | x | x | x | x (if applicable) |
| Concomitant Therapy | x | x | x | x | x | x |
| Adverse Events and Serious Adverse Events |  | x | x | x | x | x |
| <sup>1</sup> only at screening<br><sup>2</sup> including hematology, blood chemistry<br><sup>3</sup>Screening with serum pregnancy test, other visits with urine stick test<br><sup>4</sup> SIBDQ = Short Inflammatory Bowel Disease Questionnaire, Patient Diary (including stool frequency, abdominal pain and general well-being) indicated in appendix |  |  |  |  |  |  |

**Supplementary Table 3.** Indivdiually significant associations between genus abundance and faecal calprotectin concentrations prior to the intervention.

| Genus | No. samples | Slope | Lower bound | Upper bound | p | FDR |
| --- | --- | --- | --- | --- | --- | --- |
| Haemophilus | 53 | 0.259 | 0.114 | 0.405 | 0.000832 | 0.00419 |
| Parasutterella | 46 | -0.221 | -0.345 | -0.0963 | 0.000931 | 0.00419 |
| TF01-11 | 44 | 0.192 | 0.0588 | 0.326 | 0.00578 | 0.0173 |
| Hungatella_A | 42 | 0.234 | 0.0532 | 0.416 | 0.0122 | 0.024 |
| Lachnospira | 64 | 0.294 | 0.0633 | 0.524 | 0.0134 | 0.024 |
| Lawsonibacter | 65 | -0.317 | -0.602 | -0.0328 | 0.0296 | 0.0435 |
| Phascolarctobacterium | 34 | -0.135 | -0.259 | -0.011 | 0.0338 | 0.0435 |
| An114 | 54 | -0.231 | -0.45 | -0.0118 | 0.0393 | 0.0442 |
| Anaerobutyricum | 62 | -0.239 | -0.476 | -0.00159 | 0.0486 | 0.0486 |

## Supplementary Figures

**Supplementary Figure 1.**
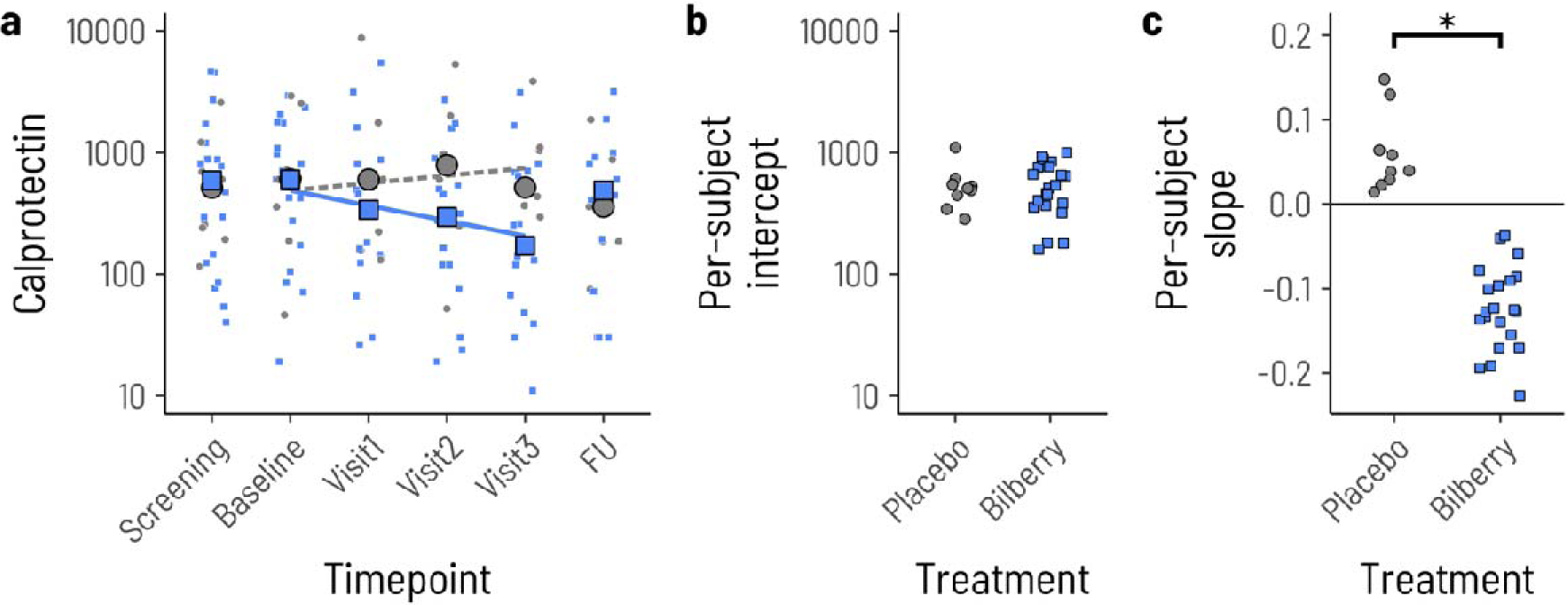
Change in the concentration of faecal calprotectin throughout the study. a. Each small point shows the concentration of faecal calprotectin (µg/g) in placebo- treated (grey) or ACRE-treated (blue) subjects at the different time points. The large circles and squares show the group geometric means. The dashed and solid lines the mean slope for placebo and ACRE, respectively, estimated from a linear mixed model with random slopes and intercepts for the two groups. b. Estimated intercept for each subject from the linear mixed model. c. Estimated slope for each subject from the linear mixed model (p = 0.036).

**Supplementary Figure 2.**
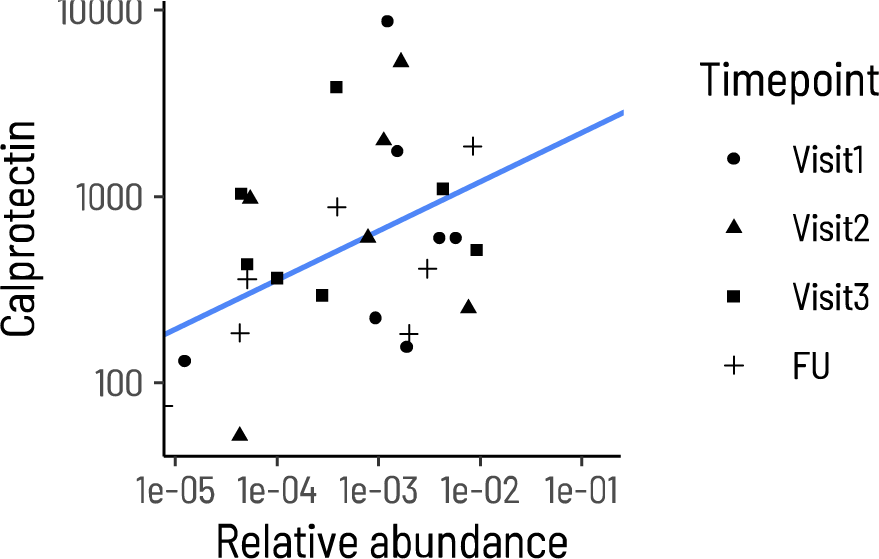
The positive correlation of Haemophilus and faecal calprotectin persists during the intervention in placebo-treated individuals. Each point corresponds to a sample from a subject at a specific timepoint, visit 1 (circles), visit 2 (triangles), visit 3 (squares), follow-up (FU, +). The line shows the estimated regression slope from a linear mixed model with subject as a random effect.

## Notes

### Clinical Trial

NCT04000139

### Funding Statement

This study was funded by the Swiss National Science Foundation (SNF) to GR [Grant No. 33IC30_166844] and the Litwin Foundation (New Hyde Park, NY)

### Author Declarations

Swiss ethics committee approval approval number: BASEC2017-00156, Date of authorisation by the ethics committee 14.02.2019 Authorisation initially for Canton Zurich (University Hospital, Prof. Rogler) and then extension of the approval to the rest participating centers (Basel, Bern, Geneva, Lausanne, St. Gallen)

## References

1 Ng SC, Shi HY, Hamidi N, et al. Worldwide incidence and prevalence of inflammatory bowel disease in the 21st century: a systematic review of population-based studies. The Lancet. 2017;390:2769–78. doi: 10.1016/S0140-6736(17)32448-0

2 Ungaro R, Mehandru S, Allen PB, et al. Ulcerative colitis. The Lancet. 2017;389:1756–70. doi: 10.1016/S0140-6736(16)32126-2

3 Schoepfer AM, Beglinger C, Straumann A, et al. Ulcerative colitis: Correlation of the Rachmilewitz Endoscopic Activity Index with fecal calprotectin, clinical activity, C-reactive protein, and blood leukocytes. Inflamm Bowel Dis. 2009;15:1851–8. doi: 10.1002/ibd.20986

4 Rogler G, Bernstein CN, Sood A, et al. Role of biological therapy for inflammatory bowel disease in developing countries. Gut. 2012;61:706–12. doi: 10.1136/gutjnl-2011-300613

5 Langhorst J, Anthonisen IB, Steder-Neukamm U, et al. Patterns of complementary and alternative medicine (CAM) use in patients with inflammatory bowel disease: Perceived stress is a potential indicator for CAM use. Complement Ther Med. 2007;15:30–7. doi: 10.1016/j.ctim.2006.03.008

6 Wing-kwan Chu, Sabrina C. M. Cheung, Roxanna A. W. Lau IFFB. Chapter 4: Bilberry (Vaccinium myrtillus L.). 2. Boca Raton: CRC Press/Taylor & Francis, Boca Raton (FL) 2011.

7 He J, Monica Giusti M. Anthocyanins: Natural colorants with health-promoting properties. Annu Rev Food Sci Technol. 2010;1:163–87. doi: 10.1146/annurev.food.080708.100754

8 Kent K, Charlton K, Roodenrys S, et al. Consumption of anthocyanin-rich cherry juice for 12 weeks improves memory and cognition in older adults with mild-to-moderate dementia. Eur J Nutr. 2017;56:333–41. doi: 10.1007/s00394-015-1083-y

9 Li S, Wang T, Fu W, et al. Role of Gut Microbiota in the Anti-Colitic Effects of Anthocyanin-Containing Potatoes. Mol Nutr Food Res. 2021;65:1–14. doi: 10.1002/mnfr.202100152

10 Mo J, Ni J, Zhang M, et al. Mulberry Anthocyanins Ameliorate DSS-Induced Ulcerative Colitis by Improving Intestinal Barrier Function and Modulating Gut Microbiota. Antioxidants. 2022;11. doi: 10.3390/antiox11091674

11 Tian L, Tan Y, Chen G, et al. Metabolism of anthocyanins and consequent effects on the gut microbiota. Crit Rev Food Sci Nutr. 2019;59:982–91. doi: 10.1080/10408398.2018.1533517

12 Farombi EO, Adedara IA, Awoyemi O V., et al. Dietary protocatechuic acid ameliorates dextran sulphate sodium-induced ulcerative colitis and hepatotoxicity in rats. Food Funct. 2016;7:913–21. doi: 10.1039/c5fo01228g

13 Li J, Wu T, Li N, et al. Bilberry anthocyanin extract promotes intestinal barrier function and inhibits digestive enzyme activity by regulating the gut microbiota in aging rats. Food Funct. 2019;10:333–43. doi: 10.1039/c8fo01962b

14 Biedermann L, Mwinyi J, Scharl M, et al. Bilberry ingestion improves disease activity in mild to moderate ulcerative colitis - An open pilot study. J Crohns Colitis. 2013;7:271–9. doi: 10.1016/j.crohns.2012.07.010

15 Biedermann L, Doulberis M, Schreiner P, et al. A Multi-Center Randomized, Double- Blind, Placebo Controlled, Parallel Group, Phase IIa Study to Evaluate the Efficacy, Safety and Tolerability of an Anthocyanin Rich Extract (ACRE) in Patients with Ulcerative Colitis. medRxiv. 2024;2024.07.19.24310589. doi: 10.1101/2024.07.19.24310589

16 Review C, Communication S, Principles G. World Medical Association Declaration of Helsinki: ethical principles for medical research involving human subjects. J Am Coll Dent. 2014;81:14–8. doi: 10.1093/acprof:oso/9780199241323.003.0025

17 European Medicines Agency (EMA). Guideline Good Clinical Practice E6(R2). Committee for Human Medicinal Products. 2018;6:1–68.

18 Tan C, Wang M, Kong Y, et al. Anti-inflammatory and intestinal microbiota modulation properties of high hydrostatic pressure treated cyanidin-3-glucoside and blueberry pectin complexes on dextran sodium sulfate-induced ulcerative colitis mice. Food Funct. 2022;4384–98. doi: 10.1039/d1fo03376j

19 Moon HJ, Cha YS, Kim KA. Blackcurrant Alleviates Dextran Sulfate Sodium (DSS)- Induced Colitis in Mice. Foods. 2023;12. doi: 10.3390/foods12051073

20 Wu B, Cox AD, Chang H, et al. Maize near-isogenic lines with enhanced flavonoids alleviated dextran sodium sulfate-induced murine colitis via modulation of the gut microbiota. Food Funct. 2023;14:9606–16. doi: 10.1039/d3fo02953k

21 Gevers D, Kugathasan S, Denson LA, et al. The Treatment-Naive Microbiome in New- Onset Crohn’s Disease. Cell Host Microbe. 2014;15:382–92. doi: 10.1016/j.chom.2014.02.005

22 Lloyd-Price J, Arze C, Ananthakrishnan AN, et al. Multi-omics of the gut microbial ecosystem in inflammatory bowel diseases. Nature. 2019;569:655–62. doi: 10.1038/s41586-019-1237-9

23 Kansal S, Catto-Smith AG, Boniface K, et al. The Microbiome in Paediatric Crohn’s Disease—A Longitudinal, Prospective, Single-Centre Study. J Crohns Colitis. 2019;13:1044–54. doi: 10.1093/ecco-jcc/jjz016

24 Putignani L, Oliva S, Isoldi S, et al. Fecal and mucosal microbiota profiling in pediatric inflammatory bowel diseases. Eur J Gastroenterol Hepatol. 2021;33:1376–86. doi: 10.1097/MEG.0000000000002050

25 Schirmer M, Denson L, Vlamakis H, et al. Compositional and Temporal Changes in the Gut Microbiome of Pediatric Ulcerative Colitis Patients Are Linked to Disease Course. Cell Host Microbe. 2018;24:600–610.e4. doi: 10.1016/j.chom.2018.09.009

26 Barberio B, Facchin S, Patuzzi I, et al. A specific microbiota signature is associated to various degrees of ulcerative colitis as assessed by a machine learning approach. Gut Microbes. 2022;14:1–12. doi: 10.1080/19490976.2022.2028366

27 Čipčić Paljetak H, Barešić A, Panek M, et al. Gut microbiota in mucosa and feces of newly diagnosed, treatment-naïve adult inflammatory bowel disease and irritable bowel syndrome patients. Gut Microbes. 2022;14:1–21. doi: 10.1080/19490976.2022.2083419

28 Shapiro JM, de Zoete MR, Palm NW, et al. Immunoglobulin A Targets a Unique Subset of the Microbiota in Inflammatory Bowel Disease. Cell Host Microbe. 2021;29:83–93.e3. doi: 10.1016/j.chom.2020.12.003

29 Pabst O. New concepts in the generation and functions of IgA. Nat Rev Immunol. 2012;12:821–32. doi: 10.1038/nri3322

30 Chow J, Tang H, Mazmanian SK. Pathobionts of the gastrointestinal microbiota and inflammatory disease. Curr Opin Immunol. 2011;23:473–80. doi: 10.1016/j.coi.2011.07.010

31 Palm NW, De Zoete MR, Cullen TW, et al. Immunoglobulin A coating identifies colitogenic bacteria in inflammatory bowel disease. Cell. 2014;158:1000–10. doi: 10.1016/j.cell.2014.08.006

32 Shapiro JM, Cho JH, Sands BE, et al. Bridging the gap between host immune response and intestinal dysbiosis in inflammatory bowel disease: Does immunoglobulin a mark the spot? Clinical Gastroenterology and Hepatology. 2015;13:842–6. doi: 10.1016/j.cgh.2015.02.028

33 Lin R, Chen H, Shu W, et al. Clinical significance of soluble immunoglobulins A and G and their coated bacteria in feces of patients with inflammatory bowel disease. J Transl Med. 2018;16:1–15. doi: 10.1186/s12967-018-1723-0

34 Gupta S, Basu S, Bal V, et al. Gut IgA abundance in adult life is a major determinant of resistance to dextran sodium sulfate-colitis and can compensate for the effects of inadequate maternal IgA received by neonates. Immunology. 2019;158:19–34. doi: 10.1111/imm.13091

35 Liu X, Wang L, Zhuang H, et al. Promoting intestinal IgA production in mice by oral administration with anthocyanins. Front Immunol. 2022;13. doi: 10.3389/fimmu.2022.826597

36 Taira T, Yamaguchi S, Takahashi A, et al. Dietary polyphenols increase fecal mucin and immunoglobulin A and ameliorate the disturbance in gut microbiota caused by a high fat diet. J Clin Biochem Nutr. 2015;57:212–6. doi: 10.3164/jcbn.15-15

